# Levels and tracking of Lipoprotein (a) serum concentrations from infancy to adolescence

**DOI:** 10.64898/2026.06.25.26356629

**Authors:** Tomi Jumpponen, Markus Juonala, Pia Salo, Irina Lisinen, Tomi T. Laitinen, Katja Pahkala, Suvi Rovio, Jorma S.A. Viikari, Tapani Rönnemaa, Harri Niinikoski, Taina Routi, Antti Jula, Joel Nuotio, Olli T. Raitakari, Juha Mykkänen

**Affiliations:** Research Centre of Applied and Preventive Cardiovascular Medicine, University of Turku, Turku, Finland; Centre for Population Health Research, University of Turku and Turku University Hospital, Turku, Finland; Department of Medicine, University of Turku, Turku, Finland; Department of Clinical Physiology and Nuclear Medicine, University of Turku and Turku University Hospital, Turku, Finland; Paavo Nurmi Centre, Unit of Health and Physical Activity, University of Turku, Turku, Finland; Department of Pediatrics and Adolescent Medicine, University of Turku and Turku University Hospital, Turku, Finland; Department of Chronic Disease Prevention, National Institute for Health and Welfare, Turku, Finland; Department of Public Health, University of Turku, Turku University Hospital, Turku, Finland; Division of Medicine, Turku University Hospital, Turku, Finland; Heart Center, Turku University Hospital and University of Turku, Turku, Finland

**Keywords:** Apolipoprotein (a), Childhood, Cardiovascular, Longitudinal, Tracking, Nutrition

## Abstract

**Background and aims:** Longitudinal data and tracking of serum lipoprotein(a) (Lp(a)) concentrations through childhood’s development from infancy to adolescence are lacking. We aimed to establish the strength of the tracking phenomenon from early infancy to adolescence and to examine whether a heart-healthy dietary intervention and individual dietary components influence serum concentrations of Lp(a).

**Methods:** 1062 healthy children aged 7 months were recruited and randomized into control (N=522) and intervention (N=540) groups in the Special Turku Coronary Risk Factor Intervention Project (STRIP). Serum Lp(a) concentration was measured at 10 age points (0.7, 1.3, 2, 3, 4, 5, 9, 11, 13, and 15 years) and median serum Lp(a) levels were studied longitudinally. Tracking across age points was studied using Spearman’s rank-order correlation. Sex differences and the effects of the dietary intervention and individual dietary components were analyzed with linear mixed-effects models for repeated measures.

**Results:** A total of 7018 Lp(a) measurements were analyzed. Median serum Lp(a) concentrations increased from infancy until approximately age 13 years. After age 13, median Lp(a) declined in boys (−15.1%) but was largely unchanged in girls. Girls had higher Lp(a) concentrations at all age points (P=0.01). Spearman’s correlation analysis indicated a strong tracking between age points in both sexes (r=0.854-0.956). Achievement of at least one dietary fat quality goal of the intervention corresponded to a 2.5% increase in serum Lp(a) concentration (P=0.0004). Higher sucrose intake was associated with modestly higher Lp(a), whereas fiber intake showed no association. At age 15 years, 16.2% of all participants with available measurements had elevated Lp(a) (≥ 30mg/dL).

**Conclusions:** A rising trend was observed in median serum Lp(a) concentrations from infancy to adolescence. Due to strong tracking, these findings suggest that early-life measurements may provide valuable insight for longitudinal cardiovascular risk assessment. Heart-healthy diet does not meaningfully influence serum Lp(a).

## INTRODUCTION

Elevated serum lipoprotein(a) [Lp(a)] concentration is an established, independent and causal risk factor for atherosclerotic cardiovascular disease (ASCVD), coronary heart disease, stroke, heart failure, and calcific aortic valve stenosis(1–5). Based on per-particle analyses, Lp(a) particles are associated with an approximately 6-fold higher risk of cardiovascular events compared with non-Lp(a) apolipoprotein B100–containing lipoproteins(6). Elevated Lp(a) levels are common, with a prevalence of approximately one in four in the general adult population, although this varies by ethnicity(7,8). Elevated serum Lp(a) levels measured in childhood have been identified as an independent risk factor for future adverse cardiovascular outcomes, with concentrations ≥30 mg/dL in youth associated with an approximately two-fold higher risk of adult atherosclerotic cardiovascular disease(9).

Serum Lp(a) concentration is predominantly genetically determined, with the vast majority of inter-individual variability attributable to inherited variation in the LPA gene(10). In adults, Lp(a) is highly stable throughout life. The Young Finns Study reported a Spearman rank-order tracking coefficient of r = 0.85 over 25 years(11). In children the evidence is more limited as prior studies have examined Lp(a) over relatively short intervals in infancy(12) or beginning at school age in clinic-referred population(13). No study to date has characterized the full longitudinal trajectory of serum Lp(a) from infancy through adolescence. This is an important gap, since Lp(a) levels are low at birth and rise in the first months of life but whether and how concentrations continue to change through childhood and adolescence, and whether trajectories diverge by sex has not yet been characterized in a prospective longitudinal cohort.

Dietary factors influence serum Lp(a) concentrations, though findings across intervention studies have been inconsistent and often counterintuitive. While heart-healthy diet reduces childhood LDL-C(14), the relationship between diet and Lp(a) remains poorly characterized(15).

This study aims to establish the strength of the tracking phenomenon from infancy to adolescence, to report the levels of serum Lp(a) at key growth and developmental milestones, and to investigate whether dietary intervention and achievement of its fat-quality goals, and intake of saturated fat, sucrose and fiber influence serum Lp(a) concentrations.

## MATERIALS AND METHODS

### Study population

The Special Turku Coronary Risk Factor Intervention Project (STRIP) was launched in 1990 aiming to prevent atherosclerosis by a dietary counselling intervention that began in infancy. A total of 1062 infants aged 7 months were enrolled in the study and randomized to intervention group (N = 540) and control group (N = 522). The intervention families and participants were given individualized nutritional counselling at 3- to 12-month intervals, at the project study site. The families and participants in the control group were met biannually until age 7 years and annually thereafter. They received basic health education, but no specific preventive guidance was provided. The intervention continued until the age of 20 years though the present analysis includes follow-up to age 15 years. The primary aim of the dietary counselling was the replacement of saturated fat with unsaturated fat in the child’s diet. Additional heart-healthy dietary recommendations were given according to the Nordic nutrition guidelines. Additional details about the study design are available elsewhere(16).

### Assessment of Dietary Intake and the Main Intervention Target

Information of dietary fat intake of participants was collected by using food diaries consisting of 4 consecutive days, including at least 1 weekend day(17). Food and nutrient intakes were analyzed using Micro Nutrica program after a dietician reviewed all food records for accuracy(18). Dietary data from a total of 5806 food diaries were included in this study. Ten age points from seven months to 15 years were initially included in the current study design. The 7-month age point was excluded from dietary analyses due to considerable differences in serum Lp(a)-levels between breastfed and formula-fed infants that could not be accounted for by macronutrient composition alone. Adherence to the main dietary intervention target was defined as achieving one or both goals of dietary fat quality: saturated fatty acid intake <10 % of total energy intake and/or unsaturated to saturated fatty acid ratio of ≥2:1(19).

### Laboratory methods

Non-fasting venous blood samples were taken at 0.7, 1.3, 2, 3, 4 years of age. Fasting venous blood samples were obtained at 5, 9, 11, 13, and 15 years of age. After clotting at room temperature and low-speed centrifugation (at 3400g for 12 minutes), serum was separated and stored at −25°C for maximally two weeks and thereafter at −75°C. A solid-phase immunoradiometric assay using direct sandwich method was employed to determine serum Lp(a) levels (Pharmacia/Mercodia, Uppsala, Sweden) (20)

### Statistical analyses

Due to the skewness of Lp(a) measurement distribution, median concentrations of Lp(a) were used to describe Lp(a) levels across age points. Tracking of Lp(a) over time was assessed using Spearman’s rank-order correlation coefficients calculated between all measurement ages. To test whether tracking coefficients differed between sexes, Pearson correlation coefficients were computed on log-transformed Lp(a) separately for girls and boys and transformed using

Fisher’s r-to-z method. Differences between sexes for each correlation coefficient were tested by comparing the z-transformed coefficients using two-sided z-tests, accounting for the sample size for each correlation. Resulting *p*-values were adjusted for multiple comparisons using the Benjamini–Hochberg false discovery rate (FDR) procedure. The longitudinal analysis of Lp(a) concentrations by age and sex, as well as the analysis of dietary intervention/control group were conducted with log-transformed Lp(a) data using linear mixed-effects models for repeated measures with ante-dependence covariance structure. The relative risk of achieving dietary fat quality targets in the intervention versus control group was estimated using a log-binomial model. A linear mixed-effects model for repeated measures was used for dietary target analyses, with age treated as a categorical covariate and an unstructured covariance matrix applied. Chi-square test was employed to test the statistical significance of categorical transitions at ages 2 and 15 years between low (<30 mg/dL) and elevated (≥30 mg/dL) Lp(a) concentrations, with the 30 mg/dL threshold associated with approximately doubled risk of adult atherosclerotic cardiovascular disease when measured in youth(9). Change in Lp(a) concentration between age 13 and 15 years was assessed using paired t-test with log-transformed values and including only individuals with measurements available at both time points (boys, N=276; girls N=263). All statistical analyses were performed using SAS version 9.4 (SAS Institute Inc, Cary, NC). Results were considered statistically significant with P <0.05.

### Ethical considerations

This study was approved by the Joint commission ethics committee of Turku University and Turku University Central Hospital. De-identified participant data were used. Written informed consent was obtained from parents at study admission and from the participants at age 15 onwards. The study adhered to the principles of the Declaration of Helsinki.

## RESULTS

### Distribution of lipoprotein(a) levels according to age and sex

The STRIP study enrolled 1062 children born in 1989-1991 and assigned into intervention (N = 540) or control (N = 522) groups. The overall participation rate for the current study was high with the compliance rate being highest at age 1.3 years with total of 863 Lp(a) measurements (81.3% of study population) and lowest at age 15 years with rate of 52.3% (Supplemental Table 1). The overall participation rate remained comparable between girls and boys. The primary reasons for discontinuation were geographic relocation, recurrent illnesses, and difficulties with blood draws(16).

A total of 7018 Lp(a) concentration measurements were obtained for this study. Table 1 presents the median Lp(a) levels of the study population categorized by age and sex. Median Lp(a) levels ranged from 6.1 mg/dL (boys at age 7 months) to 11.9 mg/dL (girls at age 13 years). Girls maintained higher longitudinal Lp(a) levels (P=0.01) (Figure 1A), with the largest age-point difference at age 15 years with median sex difference being 2.9 mg/dL (P=0.0009). In both sexes, median Lp(a) was lowest at age of seven months, rose through infancy followed by a slight decrease at ages 2 and 3 years, before increasing steadily through age 13 years (Figure 1A). Linear mixed-effects modelling confirmed a significant positive association between age and serum Lp(a) concentration in both sexes (P<0.001). Between ages 2 and 15 years, median Lp(a) concentrations increased by 18% in boys and 44% in girls (Table 1). In both sexes, median Lp(a) was lower at age 15 years than at age 13 years, with a larger median decrease in boys (10.25 to 8.70 mg/dL) than in girls (11.87 to 11.60 mg/dL). Paired analysis of log-transformed values gave a significant geometric-mean decline in both sexes between these ages, larger in boys (−19.7%; 95% CI -22.7 to -16.5) than in girls (−12.9%; 95% CI -16.2 to - 9.4; P<0.0001). The change differed by sex (sex × age interaction, P=0.0028). Participants with paired measurements at age 7 months and 15 years (N=387) had a mean Lp(a) increase of 6.01 mg/dL when comparing infant levels to adolescent levels (P<0.0001; Supplemental Figure 1).

**Table 1.** Serum lipoprotein(a) levels of study population by age and sex.

| Age(years) |  |  |  |  |  |  |
| --- | --- | --- | --- | --- | --- | --- |
|  | Boys |  |  | Girls |  |  |
|  | N | Lp(a) | IQR | N | Lp(a) | IQR |
| 7 mo | 391 | 6.10 | 4.76-9.25 | 358 | 6.44 | 4.83-10.59 |
| 13 mo | 440 | 8.15 | 5.87-13.78 | 423 | 9.12 | 6.10-17.90 |
| 2 | 401 | 7.37 | 5.43-12.00 | 389 | 8.05 | 5.63-15.82 |
| 3 | 416 | 7.34 | 5.30-12.00 | 379 | 7.87 | 5.56-16.37 |
| 4 | 403 | 7.87 | 5.45-12.70 | 373 | 8.70 | 5.78-15.37 |
| 5 | 382 | 8.07 | 5.53-14.20 | 343 | 8.95 | 5.95-17.96 |
| 9 | 303 | 9.70 | 6.11-16.96 | 287 | 11.37 | 7.28-20.71 |
| 11 | 309 | 9.78 | 6.28-16.37 | 289 | 11.62 | 6.78-21.79 |
| 13 | 300 | 10.25 | 6.03-18.33 | 277 | 11.87 | 6.78-25.79 |
| 15 | 284 | 8.70 | 4.50-15.80 | 271 | 11.60 | 6.10-24.70 |

Values of Lipoprotein (a) [Lp(a)] are presented in median concentrations (mg/dL) with corresponding interquartile ranges (IQR). N indicates the number of Lp(a) measurements.

**Figure 1.**
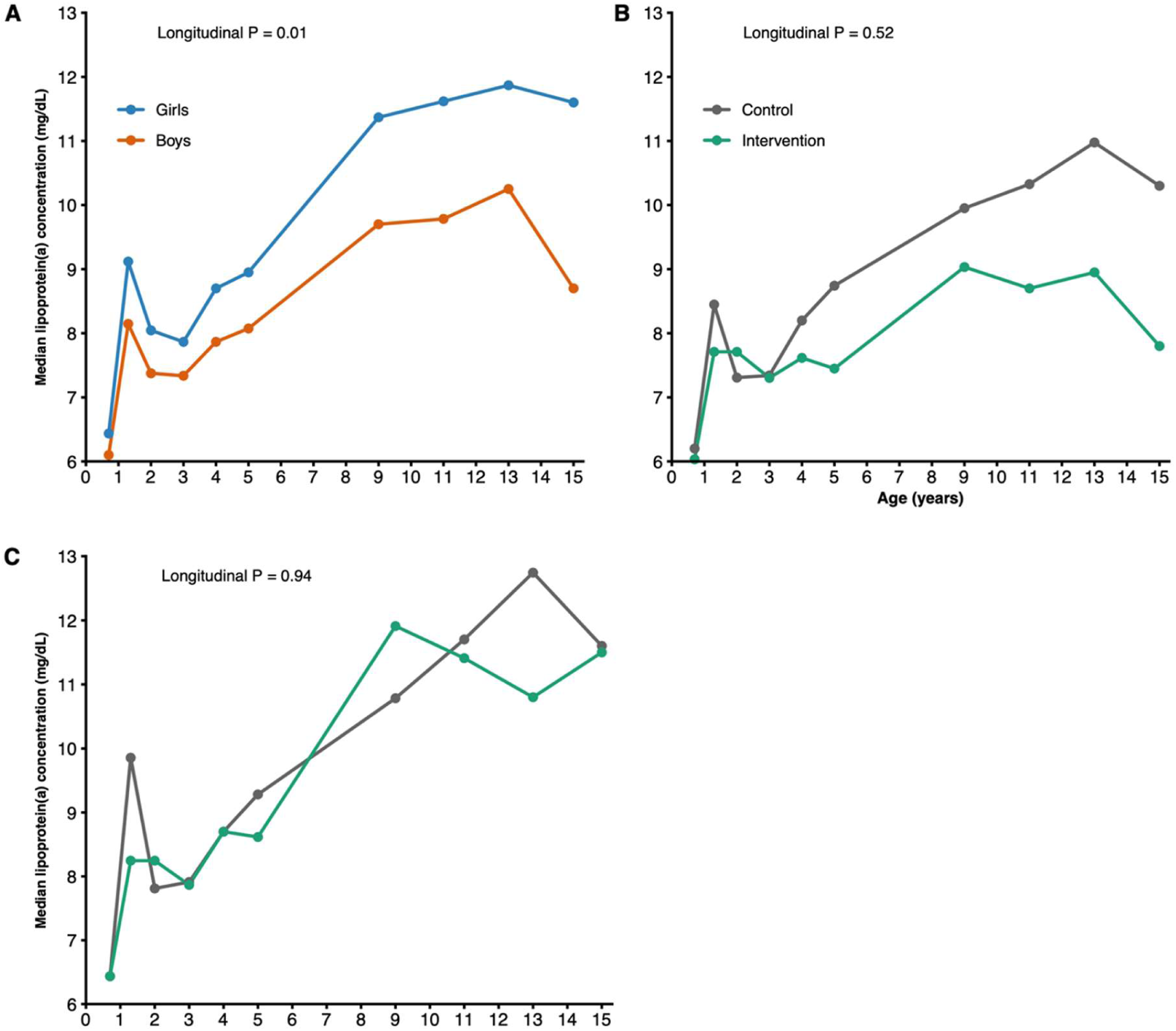
Median lipoprotein (a) concentrations (mg/dL) across 10 age points from infancy to adolescence (0.7 - 15 years). **A**) Median serum lipoprotein(a) concentrations for boys (red line) and girls (blue line). **B**) Median serum lipoprotein(a) concentrations for boys in the intervention group (green line) and control group (grey line). **C**) Median serum lipoprotein (a) concentrations for girls in the intervention group (green line) and control group (grey line). Longitudinal P-values for group differences (girls vs. boys or intervention vs. control) were derived from linear mixed-effects models for repeated measures using log-transformed Lp(a) concentrations.

### Tracking of Lp(a)

To study the effects of age and sex, Spearman’s rank order correlation coefficients were calculated separately for boys and girls (Figure 2). Correlations were consistently strong across the study years for both sexes (r = 0.854–0.956; P<0.0001 for all). Girls demonstrated slightly higher coefficients (r = 0.879–0.956; P<0.0001 for all; Figure 2A) compared to boys (r = 0.854–0.955; P< 0.0001 for all; Figure 2B). Pairwise comparison of the sex-specific coefficients showed no significant difference after Benjamini-Hochberg correction (FDR=0.43). Spearman’s rank order correlation coefficients in both sexes are presented in Supplemental Figure 2.

**Figure 2.**
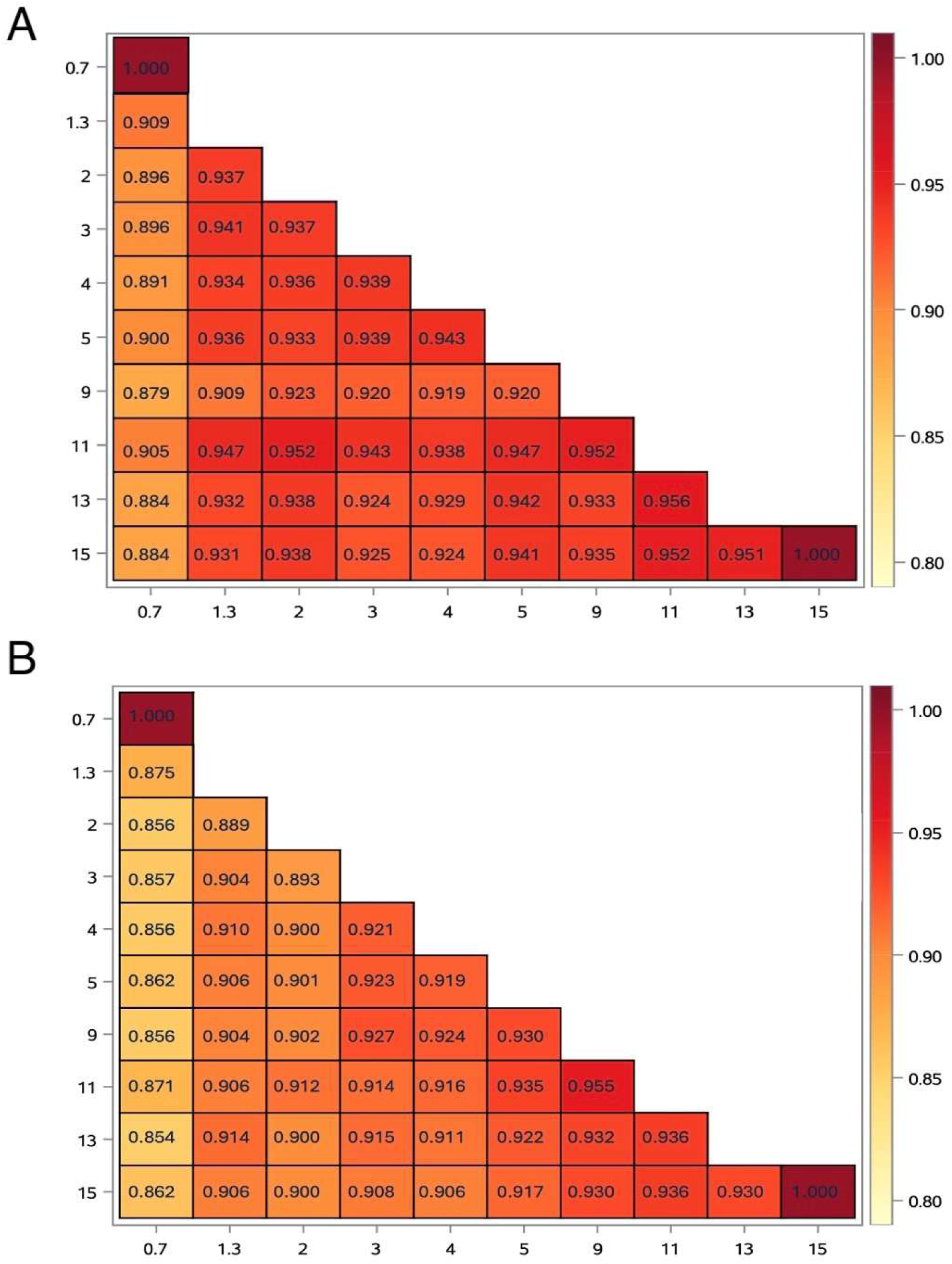
Heatmap illustration of tracking of Lp(a) levels from ages 0.7 to 15 years separately for girls (A) and boys (B). Spearman’s correlation coefficients (P<0.0001 for all) between age groups are shown in pairwise cells. Color intensity reflect correlation strength, with darker shades indicating stronger correlation.

In addition to Spearman’s rank order correlation, tracking of serum Lp(a) was assessed by cross-tabulating participants with low (<30 mg/dL) and elevated (≥30 mg/dL) Lp(a) concentration at age 2 and 15 years (Supplemental Table 2). Paired measurements were available for 469 participants. The individual-level change in Lp(a) between these two ages is shown in Supplemental Figure 3.

Of the participants with low Lp(a) levels at age 2 years, 92.7% (N=391) remained in the low category at age 15 years, while 31 (7.3%) shifted to elevated category by age 15 years (P<0.001). Of participants with elevated Lp(a) levels at age 2 years, 95.7% (N=45) remained in the elevated category. Only 2 (4.3%) participants with elevated levels at age 2 years shifted to low category by age 15 years, although Lp(a) concentrations in this marginal transition group were close to the categorical cut-off values. A total of 76 out of 469 with paired measurements (16.2%) had elevated Lp(a) levels at age 15 years. Of all available measurements at age 15 (N=555), 90 (16.2%) had elevated Lp(a) (≥30 mg/dL) and 32 (5.8%) had levels ≥50 mg/dL.

### Intervention effects and association with dietary fatty acid intake

The effects of intervention on serum Lp(a) concentration were studied separately for boys and girls (Table 2). In boys, the median Lp(a) concentration after age 3 years was consistently lower in the intervention group compared to the control group, however, the group difference was not statistically significant (Figure 1B; P=0.52). There was no notable intervention group difference seen in girls (Figure 1C; P=0.96).

**Table 2.** Serum lipoprotein(a) levels of the study population by intervention group, age and sex.

| Age<br>(years) | Intervention |  |  |  |  |  | Control |  |  |  |  |  |
| --- | --- | --- | --- | --- | --- | --- | --- | --- | --- | --- | --- | --- |
|  | Boys |  |  | Girls |  |  | Boys |  |  | Girls |  |  |
|  | N | Lp(a) | IQR | N | Lp(a) | IQR | N | Lp(a) | IQR | N | Lp(a) | IQR |
| **7 mo** | 201 | 6.03 | 4.76-8.92 | 175 | 6.44 | 4.83-11.40 | 190 | 6.20 | 4.76-9.86 | 183 | 6.44 | 4.89-10.33 |
| **13 mo** | 236 | 7.71 | 5.77-13.24 | 209 | 8.25 | 6.17-18.91 | 204 | 8.45 | 6.00-14.48 | 214 | 9.86 | 5.97-16.83 |
| **2** | 207 | 7.71 | 5.50-11.93 | 187 | 8.25 | 5.97-16.63 | 194 | 7.31 | 5.43-13.34 | 202 | 7.81 | 5.43-15.29 |
| **3** | 210 | 7.30 | 5.28-11.70 | 190 | 7.87 | 5.56-16.90 | 206 | 7.34 | 5.36-12.40 | 189 | 7.91 | 5.56-16.36 |
| **4** | 200 | 7.62 | 5.4-12.74 | 177 | 8.70 | 5.95-16.37 | 203 | 8.20 | 5.70-12.29 | 196 | 8.70 | 5.57-14.12 |
| **5** | 194 | 7.45 | 5.61-13.54 | 173 | 8.62 | 5.95-16.79 | 188 | 8.74 | 5.45-14.62 | 170 | 9.28 | 5.95-18.79 |
| **9** | 145 | 9.03 | 6.11-17.04 | 138 | 11.91 | 7.03-22.54 | 158 | 9.95 | 6.11-16.45 | 149 | 10.78 | 7.37-19.54 |
| **11** | 145 | 8.70 | 5.86-17.46 | 134 | 11.41 | 7.20-21.37 | 164 | 10.33 | 6.36-15.95 | 155 | 11.70 | 6.61-22.46 |
| **13** | 144 | 8.96 | 5.45-17.54 | 131 | 10.8 | 6.28-24.79 | 156 | 10.98 | 6.66-19.54 | 146 | 12.74 | 7.37-30.30 |
| **15** | 135 | 7.80 | 4.2-14.7 | 122 | 11.50 | 5.70-22.10 | 149 | 10.30 | 4.90-16.30 | 149 | 11.60 | 6.20-27.90 |

Values of Lipoprotein (a) [Lp(a)] are presented in median concentrations (mg/dL) with corresponding interquartile ranges (IQR). N indicates the number of Lp(a) measurements.

Participants in the intervention group achieved the targets for dietary fat quality (saturated fatty acid intake <10 % of total energy intake and/or unsaturated to saturated fatty acid ratio of ≥2:1) more often than participants in the control group. The mean proportion of participants who succeeded in achieving at least one of the dietary targets in the intervention and control groups was 34.9% vs. 9.1% (RR = 3.84; 95% CI: 3.31-4.45; P<0.0001). After adjustment for intervention group assignment, achievement of at least one dietary target corresponded to a 2.5 % increase in serum Lp(a) levels (95% CI: 1.1% - 3.9%; P=0.0004).

The analysis of dietary saturated fatty acid intake (as a proportion of total fat intake) showed that a 10% increase in saturated fatty acid intake was associated with a 1.7 % decrease in serum Lp(a) concentration (95% CI: -2.7 to -0.6 %; P=0.0023).

Associations of dietary sucrose and fiber intake with serum Lp(a) were examined by comparing participants in the highest quintile of intake (≥80th percentile) with the remaining participants, adjusted for intervention group assignment. Participants in the highest quintile of sucrose intake had 2.1% higher serum Lp(a) concentration than participants below the 80th percentile (95% CI: 0.85 to 3.3%; P=0.0011). Fiber intake in the highest quintile was not associated with serum Lp(a) concentration (0.009%; 95% CI: -1.3 to 1.3%; P=0.99).

## DISCUSSION

This longitudinal study analyzed the development of serum Lp(a) levels from infancy to adolescence. We showed that serum Lp(a) concentrations increased from infancy to approximately age 13 years and demonstrated strong tracking across childhood, with girls maintaining higher concentrations than boys at all age points. Achievement of intervention target for dietary fat quality was associated with higher serum Lp(a) levels, while the dietary intervention in itself had no effect on serum Lp(a) concentration. At age 15 years, 16.2% of all participants had elevated Lp(a) concentrations (≥30 mg/dL), and 7.3% of those with low Lp(a) levels measured at age 2 years had elevated Lp(a) levels (≥30 mg/dL) by age 15 years.

The trajectory we describe both confirms and refines the existing pediatric literature. The COMPARE study established that Lp(a) rises rapidly in the first months of life from very low cord-blood concentrations(12). The 2022 European Atherosclerosis Society consensus statement acknowledged that Lp(a) levels may continue to rise until adulthood and that multiple testing may be required in children (21). Direct evidence quantifying this post-infancy rise in a single longitudinal cohort has been lacking. Our data show that median Lp(a) continues to rise after age 2. Between ages 2 and 13 years, median Lp(a) increased by approximately 39% in boys and 47% in girls. The developmental trajectory of Lp(a) extends well beyond infancy. The biological mechanism underlying the continued rise of Lp(a) through childhood is not established, but possible contributors include developmentally regulated transcription at the LPA locus, changes in clearance, hormonal changes, environmental factors, and other unknown factors (22). The persistently higher concentrations in girls are consistent with the adult sex difference, in which women have higher Lp(a) than age-matched men (23).

Spearman rank-order coefficients between 0.85 and 0.96 across all pairwise age comparisons indicate that an individual’s relative position in the Lp(a) distribution is established early in childhood and largely preserved through adolescence. The degree of tracking is comparable to that reported in the Young Finns Study, in which the 25-year Spearman coefficient was 0.85 (11). The rank-order stability we observed in children is similar to that documented in adulthood, despite the substantial rise in absolute concentrations across adolescence. The strong tracking makes early-life Lp(a) measurement informative, particularly for identifying children with persistently elevated concentrations. Children with elevated Lp(a) in early life largely remained in the elevated category through adolescence. However, 7.3% of children with low levels at age 2 years crossed the categorical threshold into the elevated category by age 15 years, indicating that a single very early measurement does not exclude later elevation. These observations support measurement of Lp(a) at least once in childhood, with consideration of repeat measurement later in life for children whose early-life concentrations are near the categorical threshold or whose family history indicates elevated cardiovascular risk.

In boys, the Lp(a) concentration declined after age 13 years from a median of 10.25 mg/dL to 8.70 mg/dL, corresponding to a 15.1% decrease. This decline may be explained by changes in testosterone levels, as previous studies have reported an Lp(a)-lowering effect of exogenous testosterone administration of approximately 28 to 37% (24), and the rapid rise in endogenous testosterone during puberty in boys provides a plausible mechanism. The absence of a comparable median change in girls reflects a decline concentrated among those with higher concentrations, leaving the median largely unaffected. Adult women have higher Lp(a) than men, with a further rise after the menopause(23), and post-menopausal hormone therapy lowers Lp(a)(25). Together these observations suggest a complex relationship between endogenous and exogenous estradiol that warrants further investigation. The pubertal decline in boys does not undermine the tracking conclusion, because the rank order of Lp(a) values is largely preserved across this transition.

The randomized intervention did not significantly alter Lp(a) in either sex, despite producing a nearly four-fold increase in achievement of dietary fat quality targets in the intervention group. That Lp(a) can be substantially modified by nutritional exposure has been demonstrated in infancy, where breastfed infants in this cohort had approximately 45% lower Lp(a) concentrations than formula-fed infants at age 7 months, with the difference disappearing after weaning (26). The mechanism of this transient effect is not established and is not explained by differences in macronutrient composition. Beyond infancy, the dietary exposures targeted by heart-healthy guidance do not appear to meaningfully move serum Lp(a) concentrations. The 2.5% increase in Lp(a) after the achievement of dietary quality target, and the 1.7% decrease per 10% increase in saturated fat as a proportion of total fat intake, are broadly consistent with the direction described in adult feeding trials (27,28) and quantified in a recent meta-analysis (29), in which lower saturated fat intake has been associated with modestly higher Lp(a), in the opposite direction to the low-density lipoprotein cholesterol response. The mechanism underlying this opposing response is not fully established but may involve competitive clearance at shared lipoprotein receptors or changes in the availability of apoB-containing particles that recycle apo(a) in the postprandial state(30). Higher sucrose intake was associated with modestly higher Lp(a) (2.1%) and fiber intake showed no association. The magnitudes observed are small and should not be read as evidence that heart-healthy diets meaningfully raise Lp(a).

This study has several limitations. Although the control group children did not receive intensive dietary or lifestyle guidance, the awareness of healthy lifestyle choices may have been prominent across this cohort compared to their age-matched peers due to consistent participation in the study protocol. This increased awareness may have attenuated the differences between groups. As a result, largely genetically driven factors such as serum Lp(a) concentrations might appear less distinct between groups. Lp(a) was measured with a mass-based immunoradiometric assay reporting concentrations in mg/dL, whereas current consensus favors isoform-insensitive molar measurement in nmol/L(21). As apoprotein(a) size varies between individuals with the number of kringle IV type 2 repeats, the mass-based assay can over- or underestimate particle number depending on a person’s isoform. This inter-individual variability may affect our cross-sectional comparisons, namely the threshold-based prevalence estimates, which should be taken into account when interpreting the results. The immunoradiometric assay detects apolipoprotein(a), which is present in a single copy per Lp(a) particle and is largely genetically determined by the LPA locus. Any isoform-related bias in mass-based measurement is expected to be similar within an individual across timepoints and therefore to affect within-person comparisons less than between-person ones. Concordantly, the observed high rank-order tracking would not be expected if measurement bias varied substantially within individuals over time. The cohort is ethnically homogeneous and Finnish, which limits generalizability given the known variation in Lp(a) distributions across ethnicities (31). Despite these limitations, the strong rank-order and categorical tracking of Lp(a) we observed across childhood supports current European and American recommendations that Lp(a) be measured at least once for cardiovascular risk stratification (32,33).

In summary, we observed an increasing trend in serum Lp(a) concentrations from infancy to adolescence in both sexes with a high degree of tracking. Girls had consistently higher Lp(a) concentrations at all age points. Better dietary fat quality was associated with a small (2.5%) increase in serum Lp(a), while intervention as such was insufficient to induce significant changes. A total of 16.2% of participants had elevated (≥30 mg/dL) serum Lp(a) concentrations at age 15. Due to the strong tracking, our findings suggest that early-life measurements may provide valuable insight for future cardiovascular risk assessment.

## AUTHOR CONTRIBUTIONS

TJ, MJ and JM conceptualized the paper. IL, KP, SR, JV, TR, HN, TR and OR collected and managed data. TJ drafted the manuscript and performed statistical analyses. IL assisted with the analyses. All authors reviewed, edited the manuscript for critical intellectual content and approved the final manuscript.

## Data Availability

Data sharing outside the STRIP group requires a data sharing agreement. Investigators can submit an expression of interest to the STRIP Steering Committee (https://stripstudy.utu.fi/en/strip-study/).

## ACKNOWLEDGEMENTS

We thank the STRIP study children, parents and grandparents for their time, effort and longstanding commitment to the project. We also acknowledge the essential efforts of clinical and administrative staff, whose contributions were critical to making this study possible.

## REFERENCES

1. Girard AS, Paulin A, Manikpurage HD, Lajeunesse E, Clavel M, Pibarot P, et al. Impact of Lipoprotein(a) on Valvular and Cardiovascular Outcomes in Patients With Calcific Aortic Valve Stenosis. JAHA. 2025 Mar 13;e038955. doi:10.1161/JAHA.124.038955

2. Clarke R, Wright N, Lin K, Yu C, Walters RG, Lv J, et al. Causal Relevance of Lp(a) for Coronary Heart Disease and Stroke Types in East Asian and European Ancestry Populations: A Mendelian Randomization Study. Circulation. 2025 Apr 29;CIRCULATIONAHA.124.072086. doi:10.1161/CIRCULATIONAHA.124.072086

3. Ballantyne CM. Lipoprotein(a) and Heart Failure. JACC: Heart Failure. 2016 Jan;4(1):88–9. doi:10.1016/j.jchf.2015.11.001

4. Singh S, Baars DP, Aggarwal K, Desai R, Singh D, Pinto-Sietsma SJ. Association between lipoprotein (a) and risk of heart failure: A systematic review and meta-analysis of Mendelian randomization studies. Current Problems in Cardiology. 2024 Apr;49(4):102439. doi:10.1016/j.cpcardiol.2024.102439

5. Nordestgaard BG, Langsted A. Lipoprotein(a) and cardiovascular disease. The Lancet. 2024 Sep;404(10459):1255–64. doi:10.1016/S0140-6736(24)01308-4

6. Björnson E, Adiels M, Taskinen MR, Burgess S, Chapman MJ, Packard CJ, et al. Lipoprotein(a) Is Markedly More Atherogenic Than LDL. Journal of the American College of Cardiology. 2024 Jan;83(3):385–95. doi:10.1016/j.jacc.2023.10.039

7. Varvel S, McConnell JP, Tsimikas S. Prevalence of Elevated Lp(a) Mass Levels and Patient Thresholds in 532 359 Patients in the United States. ATVB. 2016 Nov;36(11):2239–45. doi:10.1161/atvbaha.116.308011

8. Reyes-Soffer G, Ginsberg HN, Berglund L, Duell PB, Heffron SP, Kamstrup PR, et al. Lipoprotein(a): A Genetically Determined, Causal, and Prevalent Risk Factor for Atherosclerotic Cardiovascular Disease: A Scientific Statement From the American Heart Association. ATVB. 2022 Jan;42(1). doi:10.1161/ATV.0000000000000147

9. Raitakari O, Kartiosuo N, Pahkala K, Hutri-Kähönen N, Bazzano LA, Chen W, et al. Lipoprotein(a) in Youth and Prediction of Major Cardiovascular Outcomes in Adulthood. Circulation. 2023 Jan 3;147(1):23–31. doi:10.1161/CIRCULATIONAHA.122.060667

10. Kronenberg F, Utermann G. Lipoprotein(a): resurrected by genetics. J Intern Med. 2013 Jan;273(1):6–30. doi:10.1111/j.1365-2796.2012.02592.x

11. Raitakari O, Kivelä A, Pahkala K, Rovio S, Mykkänen J, Ahola-Olli A, et al. Long-term tracking and population characteristics of lipoprotein (a) in the Cardiovascular Risk in Young Finns Study. Atherosclerosis. 2022 Sep;356:18–27. doi:10.1016/j.atherosclerosis.2022.07.009

12. Strandkjær N, Hansen MK, Nielsen ST, Frikke-Schmidt R, Tybjærg-Hansen A, Nordestgaard BG, et al. Lipoprotein(a) Levels at Birth and in Early Childhood: The COMPARE Study. The Journal of Clinical Endocrinology & Metabolism. 2022 Jan 18;107(2):324–35. doi:10.1210/clinem/dgab734

13. De Boer LM, Hof MH, Wiegman A, Stroobants AK, Kastelein JJP, Hutten BA. Lipoprotein(a) levels from childhood to adulthood: Data in nearly 3,000 children who visited a pediatric lipid clinic. Atherosclerosis. 2022 May;349:227–32. doi:10.1016/j.atherosclerosis.2022.03.004

14. Niinikoski H, Pahkala K, Ala-Korpela M, Viikari J, Rönnemaa T, Lagström H, et al. Effect of Repeated Dietary Counseling on Serum Lipoproteins From Infancy to Adulthood. Pediatrics. 2012 Mar 1;129(3):e704–13. doi:10.1542/peds.2011-1503

15. Enkhmaa B, Petersen KS, Kris-Etherton PM, Berglund L. Diet and Lp(a): Does Dietary Change Modify Residual Cardiovascular Risk Conferred by Lp(a)? Nutrients. 2020 Jul 7;12(7):2024. doi:10.3390/nu12072024

16. Simell O, Niinikoski H, Ronnemaa T, Raitakari OT, Lagstrom H, Laurinen M, et al. Cohort Profile: The STRIP Study (Special Turku Coronary Risk Factor Intervention Project), an Infancy-onset Dietary and Life-style Intervention Trial. International Journal of Epidemiology. 2009 Jun 1;38(3):650–5. doi:10.1093/ije/dyn072

17. Talvia S, Lagström H, Räsänen M, Salminen M, Räsänen L, Salo P, et al. A Randomized Intervention Since Infancy to Reduce Intake of Saturated Fat: Calorie (Energy) and Nutrient Intakes Up to the Age of 10 Years in the Special Turku Coronary Risk Factor Intervention Project. Arch Pediatr Adolesc Med. 2004 Jan 1;158(1):41. doi:10.1001/archpedi.158.1.41

18. Hakala P, Marniemi J, Knuts LR, Kumpulainen J, Tahvonen R, Plaami S. Calculated vs analysed nutrient composition of weight reduction diets. Food Chemistry. 1996 Sep;57(1):71–5. doi:10.1016/0308-8146(96)00077-5

19. Laitinen TT, Nuotio J, Rovio SP, Niinikoski H, Juonala M, Magnussen CG, et al. Dietary Fats and Atherosclerosis From Childhood to Adulthood. Pediatrics. 2020 Apr 1;145(4):e20192786. doi:10.1542/peds.2019-2786

20. März W, Siekmeier R, Groβ E, Groβ W. Determination of lipoprotein(a): enzyme immunoassay and immunoradiometric assay compared. Clinica Chimica Acta. 1993 Feb;214(2):153–63. doi:10.1016/0009-8981(93)90107-F

21. Kronenberg F, Mora S, Stroes ESG, Ference BA, Arsenault BJ, Berglund L, et al. Lipoprotein(a) in atherosclerotic cardiovascular disease and aortic stenosis: a European Atherosclerosis Society consensus statement. European Heart Journal. 2022 Oct 14;43(39):3925–46. doi:10.1093/eurheartj/ehac361

22. Enkhmaa B, Berglund L. Non-genetic influences on lipoprotein(a) concentrations. Atherosclerosis. 2022 May;349:53–62. doi:10.1016/j.atherosclerosis.2022.04.006

23. Simony SB, Mortensen MB, Langsted A, Afzal S, Kamstrup PR, Nordestgaard BG. Sex differences of lipoprotein(a) levels and associated risk of morbidity and mortality by age: The Copenhagen General Population Study. Atherosclerosis. 2022 Aug;355:76–82. doi:10.1016/j.atherosclerosis.2022.06.1023

24. Zmuda JM, Thompson PD, Dickenson R, Bausserman LL. Testosterone decreases lipoprotein(a) in men. The American Journal of Cardiology. 1996 Jun;77(14):1244–7. doi:10.1016/S0002-9149(96)00174-9

25. Nie G, Yang X, Wang Y, Liang W, Li X, Luo Q, et al. The Effects of Menopause Hormone Therapy on Lipid Profile in Postmenopausal Women: A Systematic Review and Meta-Analysis. Front Pharmacol. 2022 Apr 12;13:850815. doi:10.3389/fphar.2022.850815

26. Routi T, Rönnemaa T, Lapinleimu H, Salo P, Viikari J, Leino A, et al. Effect of Weaning on Serum Lipoprotein(a) Concentration: The STRIP Baby Study. Pediatr Res. 1995 Oct;38(4):522–7. doi:10.1203/00006450-199510000-00008

27. Ginsberg HN, Kris-Etherton P, Dennis B, Elmer PJ, Ershow A, Lefevre M, et al. Effects of Reducing Dietary Saturated Fatty Acids on Plasma Lipids and Lipoproteins in Healthy Subjects: The Delta Study, Protocol 1. ATVB. 1998 Mar;18(3):441–9. doi:10.1161/01.ATV.18.3.441

28. Ebbeling CB, Knapp A, Johnson A, Wong JM, Greco KF, Ma C, et al. Effects of a low-carbohydrate diet on insulin-resistant dyslipoproteinemia—a randomized controlled feeding trial. The American Journal of Clinical Nutrition. 2022 Jan;115(1):154–62. doi:10.1093/ajcn/nqab287

29. Riley TM, Sapp PA, Kris-Etherton PM, Petersen KS. Effects of saturated fatty acid consumption on lipoprotein (a): a systematic review and meta-analysis of randomized controlled trials. The American Journal of Clinical Nutrition. 2024 Sep;120(3):619–29. doi:10.1016/j.ajcnut.2024.06.019

30. Kris-Etherton PM, Riley TM, Petersen KS. Dietary modulation of Lp(a): more questions than answers. Journal of Lipid Research. 2024 Aug;65(8):100592. doi:10.1016/j.jlr.2024.100592

31. Enkhmaa B, Anuurad E, Berglund L. Lipoprotein (a): impact by ethnicity and environmental and medical conditions. Journal of Lipid Research. 2016 Jul;57(7):1111– 25. doi:10.1194/jlr.R051904

32. Mach F, Koskinas KC, Roeters Van Lennep JE, Tokgözoğlu L, Badimon L, Baigent C, et al. 2025 Focused Update of the 2019 ESC/EAS Guidelines for the management of dyslipidaemias. European Heart Journal. 2025 Aug 29;ehaf190. doi:10.1093/eurheartj/ehaf190

33. Writing Committee Members, Blumenthal RS, Morris PB, Gaudino M, Johnson HM, Anderson TS, et al. 2026 ACC/AHA/AACVPR/ABC/ACPM/ADA/AGS/APhA/ASPC/NLA/PCNA Guideline on the Management of Dyslipidemia: A Report of the American College of Cardiology/American Heart Association Joint Committee on Clinical Practice Guidelines. Circulation. 2026 Mar 13;CIR.0000000000001423. doi:10.1161/CIR.0000000000001423

